# Exogenous ketosis attenuates acute mountain sickness and mitigates high-altitude hypoxemia

**DOI:** 10.1101/2023.11.21.23298762

**Authors:** Myrthe Stalmans, Domen Tominec, Wout Lauriks, Ruben Robberechts, Tadej Debevec, Chiel Poffé

**Author notes:** **Correspondence:** Chiel Poffé, Exercise Physiology Research Group, Department of Movement Sciences, KU Leuven, Tervuursevest 101, Heverlee, 3001, Belgium.

## Abstract

**Background:** Acute mountain sickness (AMS) represents a considerable issue for individuals sojourning to high altitudes with systemic hypoxemia known to be intimately involved in its development. Based on recent evidence that ketone ester (KE) intake attenuates hypoxemia, we sought to investigate whether exogenous ketosis might mitigate AMS development and improve hypoxic tolerance.

**Methods:** Fourteen healthy, male participants were enrolled in two 29h protocols (simulated altitude of 4,000-4,500m) receiving either KE or a placebo (CON) at regular timepoints throughout the protocol in a randomized, crossover manner. Select physiological responses were characterized after 15min and 4h in hypoxia, and the protocol was terminated prematurely upon development of severe AMS.

**Results:** All participants tolerated the protocol equally long (n=6, of which n=5 completed the protocol in both conditions) or longer (n=8) in KE. Overall protocol duration increased by 32% on average with KE, and doubled for AMS-developing participants. KE consistently induced diurnal ketosis, a mild metabolic acidosis, hyperventilation, and relative sympathetic dominance. KE also fully negated the progressive hypoxemia that was observed between 15min and 4h in hypoxia in CON, while concomitantly increasing cerebral oxygenation and capillary pO_2_ within this timeframe. This coincided with a KE-induced reduction in cerebral oxygen supply, suggesting that KE reduced cerebral oxygen consumption under hypoxic conditions.

**Conclusions:** These data indicate that exogenous ketosis improves hypoxic tolerance in humans and attenuates AMS development. The key underlying mechanisms include improved arterial and cerebral oxygenation, in combination with lowered cerebral blood flow and oxygen demand, and increased sympathetic dominance.

**Summary:** Ketone ester intake attenuated the development of acute mountain sickness at a simulated altitude of 4,000-4,500m. This likely resulted from a mitigation of arterial and cerebral hypoxemia, reduced cerebral blood flow and increased sympathetic drive.

## INTRODUCTION

Acute mountain sickness (AMS) is the most prevalent form of altitude illness and develops in ∼25% of individuals upon exposure to 2,500m altitude [1] with its prevalence further increasing at higher altitudes [2, 3]. It presents as a syndrome of nonspecific symptoms including headache, nausea, vomiting, dizziness or fatigue, eventually also compromising general functioning [1]. Although AMS is typically self-limiting and usually resolves within 2-3 days [4], it can progress into life-threatening conditions such as high-altitude pulmonary (HAPE) or cerebral (HACE) oedema [5]. Hypoxemia is considered to be the primary factor driving AMS pathogenesis [6], and for instance increases cerebral blood flow in order to preserve oxygen delivery to the brain which in turn may increase intracranial pressure [7, 8]. Also a hypoxemia-induced initial relative dominance of the parasympathetic nervous system was found to play a crucial role in AMS development [9]. Some pharmacological interventions (*e.g.,* acetazolamide) have been shown to reduce AMS symptoms [10, 11]. However, a recent Cochrane review concluded that there is currently no clear evidence for any non-pharmacological intervention to robustly mitigate AMS [12].

Interestingly, early studies in rodents observed that increasing ketone bodies (KB) in the blood improved tolerance to extreme hypoxia (F_I_O_2_: 4-5%) [13, 14]. KB, especially acetoacetate (AcAc) and D-β-hydroxybutyrate (βHB), are fatty acid derived molecules that are primarily produced in the liver upon conditions of energetic stress, and are preferentially utilized as an energy source by the brain under hypoxic stress [15]. More recently, we observed that increasing blood ketone bodies via ketone ester (KE) ingestion also attenuated the drop in oxygen saturation in human participants during exercise at ∼2,500-3,000m simulated altitude [16]. This may be of primary importance given the above mentioned direct relationship between the extent of hypoxemia and AMS susceptibility and severity [6].

Accordingly, these data indicate that increasing blood KB may be a useful strategy to mitigate the development of AMS. Against this background, we aimed to identify whether increasing blood KB via oral KE ingestion can inhibit the development and severity of AMS, as well as to determine the potential underlying physiological mechanism(s).

## METHODS

### Study design and participants

This randomized, double-blind, placebo-controlled, crossover study was approved by the Ethics Committee Research UZ/KU Leuven (B3222022000810), preregistered at www.clinicaltrials.gov (NCT05588427) and conducted in accordance with the Declaration of Helsinki guidelines. We recruited healthy, male participants with the following inclusion criteria: (i) age: 18-35 years, (ii) body mass index (BMI): 18-25 kg.m^-^², (iii) regular physically active: 2-7 h.week^-1^. Exclusion criteria included smoking, a history of altitude-sensitive pathologies, and pre-exposure to altitudes above 1,500m in the three months preceding the study. All participants provided written informed consent before inclusion in the study.

As AMS typically peaks after 16-24h of hypoxic exposure [4], we designed a 29h protocol (Fig. 1) in a normobaric facility with the explicit purpose to elicit AMS (see ‘Supplementary information’ for full details). Throughout both experimental sessions, separated by a 1-week washout period, participants received either ketone ester (KE) or placebo drinks (CON) in a counterbalanced order. During the first 26h of each session, the participants resided at a simulated altitude of 4,000m (F_I_O_2_: 12.7%), after which they were immediately transferred to 4,500m (F_I_O_2_: 11.8%). Throughout the protocol, participants regularly performed exercise bouts on a calibrated cycling ergometer (Avantronic Cyclus II, Leipzig, Germany) to simulate the workload associated with normal ascend rates [17] and to facilitate AMS development [18] (see ‘Supplementary information’). The protocol was terminated prematurely if the participant developed severe AMS [Lake Louise Score (LLS) of ≥ 10 out of 15 [1]], resulting in a distribution of all participants (AMS_all_) over AMS-sensitive participants that developed severe AMS in at least one of both sessions (AMS_high_), and participants that completed both sessions (AMS_low_). Both experimental sessions started on the same time of the day within a given individual (between 7:00AM and 10:00AM). The participants’ tolerated duration of the protocol (*e.g.* an index for hypoxic tolerance) was defined as the primary outcome of the study.

**Figure 1.**
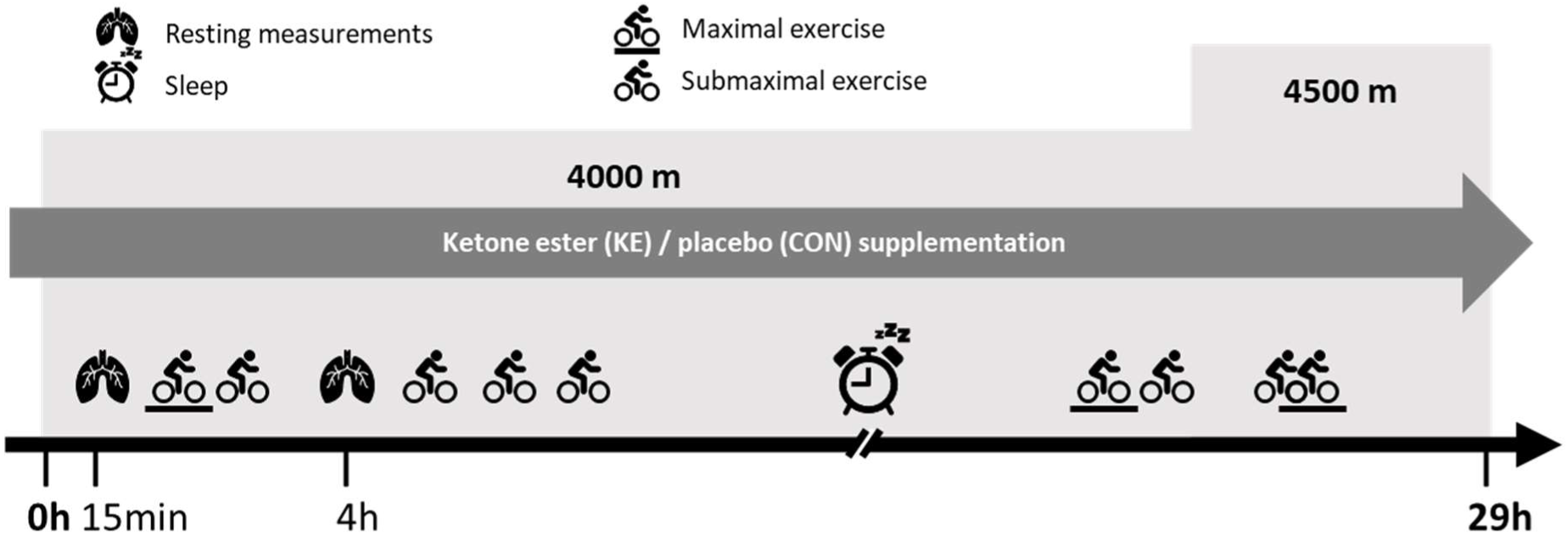
Schematic representation of the experimental sessions. In a double-blind, randomized, crossover design, 14 participants underwent a 29h normobaric hypoxic protocol (26h at 4,000m, followed by 3h at 4,500m) that was explicitly designed to elicit acute mountain sickness (AMS). Participants were instructed to comply to the protocol, but it was prematurely terminated if they developed severe AMS (Lake Louise score ≥ 10 out of 15). Throughout the entire protocol, participants received either ketone ester (KE) or placebo (CON) supplements at regular timepoints. In each session, 2 maximal (10min warming up + ramp protocol), 4 submaximal (30min) and 1 combined exercise bout (40min + ramp protocol) were completed. Moreover, after 15min and 4h in hypoxia, the following resting measurements were performed in order to characterize the physiological response: blood oxygen saturation, cerebral and muscular oxygenation status, cerebral blood flow, respiratory gas exchange, heart rate, heart rate variability, and blood pressure. In addition, before hypoxic entry, as well as after 1.5 and 3h in hypoxia, a capillary blood sample was collected for determination of acid-base balance, pO_2_ and pCO_2._

### Preliminary testing

One week before the first experimental session, participants engaged in a normoxic familiarization session in which they performed an incremental cycling test (see ‘Supplementary information’) to determine their maximal oxygen consumption rate (V̇O_2_max, *e.g.,* highest value over a 15sec period) using indirect calorimetry (Cortex Metalyzer IIIb, Leipzig, Germany). After 15min of passive recovery, participants were familiarized with all the measurements that were performed during the experimental sessions.

### Nutritional intervention

In the KE condition, participants received the ketone monoester (R)-3-hydroxybutyl (R)-3-hydroxybutyrate (KetoneAid Inc., Falls Church, Virginia, USA; ∼4.7 kcal.g^-1^) in boluses of either 25g (30min before entering hypoxia, as well as after 1, 4, 23 and 24.5h in hypoxia and 30min before sleep) or 12.5g (after 2.5, 5, 6.5, 7.5, 9, 26 and 27.5h in hypoxia) with the purpose to establish a stable ketosis ([ßHB] ∼2-5 mM) during the diurnal parts of the protocol. In CON, participants received a non-isocaloric, taste and viscosity matched placebo consisting of 12.5% w/v collagen (6d Sports Nutrition, Oudenaarde, Belgium) and 1 mM bitter sucrose octaacetate (Sigma-Aldrich, Bornem, Belgium) dissolved in water to generate either supplements of 12.5g or 25g. The bitter taste of both supplements was softened by adding 5% w/v sucralose (MyProtein, New York City, US) and 1.0% v/v strawberry flavor drops (MyProtein, New York City, US). If participants completed the full protocol, total caloric intake via the KE supplements was ∼1000 kcal *vs.* ∼100 kcal for the CON supplements. Supplements were provided in nontransparent 50 mL tubes to avoid potential visual identification. In addition, total energy intake was standardized (∼4000 kcal during the 29h protocol excluding KE/CON supplements), while water intake was *ad libitum* but replicated within a given individual, from the evening before until the end of each 29h experimental session (see ‘Supplementary information’).

### Physiological measurements

To characterize the resting physiological responses, measurement bouts were performed after 15min and 4h of hypoxic exposure. This included sequential determination of (i) blood oxygen saturation (SpO_2_) (Nellcor PM10N, Medtronic, Minneapolis, USA), (ii) cerebral (prefrontal cortex) and skeletal muscle (*m. vastus lateralis*) oxygenation status (NIRO-200, Hamamatsu, Shizuoka, Japan), (iii) cerebral blood flow (Vivid E9, EG Healthcare, New York, USA) through the internal carotid artery (ICA) and the vertebral artery (VA), which constitute the main intracranial blood supply [19] and, concurrently, oxygen delivery calculated as (VA+ICA)*SpO_2_ [19], (iv) respiratory gas exchange (Cortex Metalyzer IIIb, Cortex Medical, Leipzig, Germany), (v) heart rate and heart rate variability (HRV) (H10 chest strap, Polar, Kempele, Finland) as a measure of autonomic nervous system balance, and (vi) blood pressure (Omron M6, Omron healthcare, Kyoto, Japan), with the participants lying in supine position in bed and their eyes closed yet awake. Furthermore, capillary blood samples were obtained at regular timepoints for immediate determination of ßHB (GlucoMen Areo 2K-meter with ß-ketone sensor strips, A. Menarini Diagnostics, Firenze, Italy). We also looked at acid-base balance and capillary blood gasses which were measured before hypoxic entry and after 1.5 and 3h in hypoxia (ABL90 FLEX analyzer, Radiometer Medical ApS, Copenhagen, Denmark).In addition, we collected a venous blood sample before hypoxic entry and after 5h in hypoxia, which was used for determination of plasma glucagon (see ‘Supplementary information’). LLS was evaluated after 4h. Additional details on these methods are provided in the supplementary material.

### Statistical analyses and sample size calculation

All statistical analyses were performed in GraphPad Prism version 9.3.1 (GraphPad Software, La Jolla, CA, USA). A nonparametric Wilcoxon matched-pairs signed rank test was used to evaluate the duration of participants’ ability to comply to the hypoxic protocol (not normally distributed, D’Agostino Pearson normality test) and for LLS after 4h (discrete variable). A two-way repeated measures analysis of variance (ANOVA) was used to evaluate differences between KE and CON and over time for measurements that were performed at multiple timepoints. When a significant interaction effect was identified, post-hoc analyses were performed using Šidák’s correction. When applicable, reported p-values refer to these post-hoc analyses and otherwise, p-values for main effects were included. All data are presented as mean ± SD and statistical significance was defined as p < 0.05. Due to the absence of information regarding the effect of ketosis on hypoxic tolerance in humans, an earlier study reporting increased hypoxic tolerance in animals upon ketosis was used for the a-priori sample size calculation [13]. This indicated that a sample size of 14 per condition was required to establish a difference between both conditions (effect size dz: 0.57; α error: 0.05; power: 0.80; two-sided; Wilcoxon signed-rank test for matched pairs).

## RESULTS

### Characteristics of participants

Sixteen healthy, male participants were initially enrolled in the study with two drop-outs due to Covid-19 infection. Hence, final data analyses reported in the present paper were conducted on fourteen participants (Table 1).

**Table 1.** Participants’ baseline characteristics.

|  |  |
| --- | --- |
| Age (yr) | 26 ± 5 |
| Height (m) | 1.84 ± 0.05 |
| Body mass (kg) | 73.9 ± 4.0 |
| VO<sub>2</sub>max (mL.kg<sup>-1</sup>.min<sup>-1</sup>) | 52.0 ± 6.7 |

Data are means ± SD and represent baseline characteristics of the 14 included participants.

### KE induced ketosis, increased hypoxic tolerance and reduced AMS symptoms

The primary outcome of the study was the tolerated time in the hypoxic protocol (*e.g.,* time until severe AMS developed). From the fourteen participants (AMS_all_), five participants (36%, AMS_low_) completed the entire protocol in both conditions. From the other nine participants (AMS_high_), two completed the protocol in KE only, while seven were unable to complete the protocol in either condition but tolerated the protocol equally long or longer in KE *vs.* CON. Hence, upon KE, the tolerated duration increased by 32% when considering all participants (∼20h in KE *vs.* 15h in CON) and doubled when considering only AMS_high_ (∼15h in KE *vs.* 7.5h in CON) (Fig. 2a). After 4h in hypoxia, AMS symptoms were already evident in AMS_high_ and were, within this subgroup, more pronounced in CON *vs.* KE (Fig. 2b). Throughout the diurnal parts of the protocol, KE ingestion induced a stable ketosis (*e.g.,* blood [ßHB] of ∼3 mM), whereas blood [ßHB] remained low in CON (Fig. 3). Moreover, plasma glucagon levels increased by ∼25% in both groups after the first 5h in the hypoxic protocol (Fig. S1).

**Figure 2.**
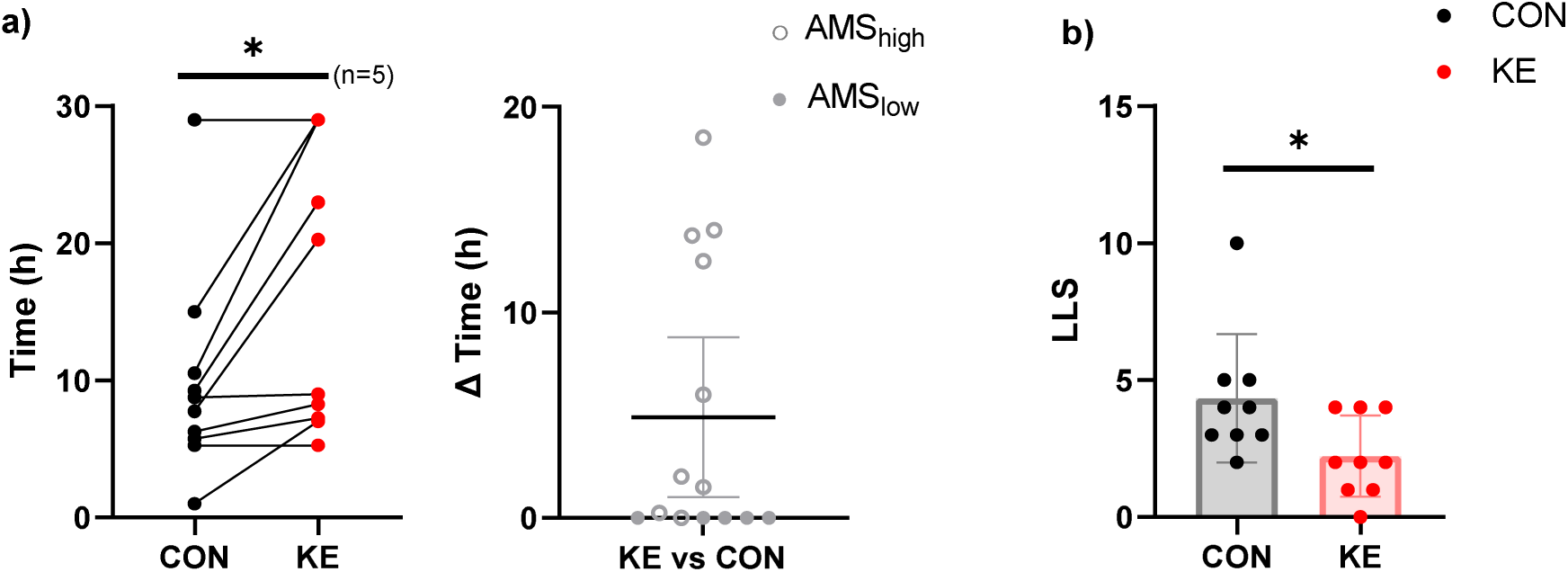
(a) Left panel: individual data points showing participants’ tolerated time in hypoxia in response to ingestion of ketone ester (KE, red) *vs.* placebo (CON, black) during a 29h normobaric hypoxic protocol. Right panel: individual differences together with mean ± 95% confidence interval for tolerated time in hypoxia between the KE and CON session, with a distinction between participants that developed severe AMS (in at least one of both sessions (AMS_high_, open circles) and participants that developed no AMS (AMS_low_, full circles). Data are shown for n = 14. (b) Mean (bar plots) ± SD (whiskers) as well as individual Lake Louise Scores (LLS) are shown after 4h in hypoxia. Only data for the participants that eventually developed AMS in at least one condition were included (n = 9). For the single participant that developed severe AMS before the 4h timepoint, we included the value that was noted down upon hypoxic leave. *, p < 0.05 between KE and CON.

**Figure 3.**
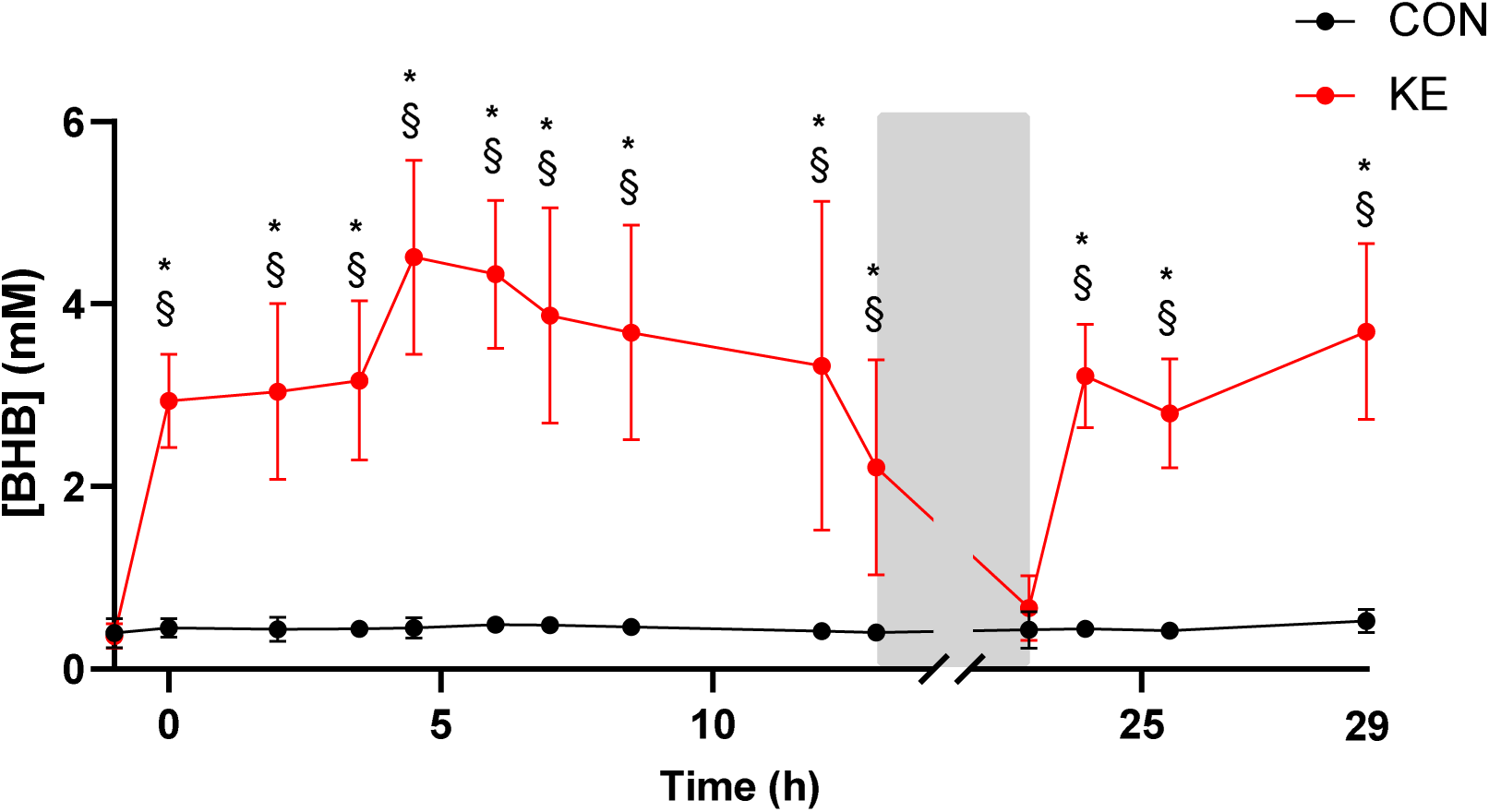
Mean ± SD data representing blood D-β-hydroxybutyrate concentration ([βHB]) in response to ingestion of ketone ester (KE, red) *vs.* placebo (CON, black) during a 29h normobaric hypoxic protocol. Grey area indicates the time during which the participants were sleeping. Data are means of all participants at the start (n = 14) however sample size decreases upon early protocol termination. *, p < 0.05 between KE and CON; §, p < 0.05 *vs.* baseline for indicated condition.

### KE mitigated hypoxemia and reduced cerebral blood flow and oxygen delivery

Participants’ SpO_2_ was comparable between KE and CON after 15min of hypoxic exposure. At the 4h timepoint, SpO_2_ had further dropped in CON by ∼5% but remained stable in KE, indicating that KE attenuated the progressive increase in hypoxemia (Fig. 4a). Interestingly, cerebral tissue oxygenation was ∼4% higher in KE *vs.* CON after 4h (Fig. 4b), while blood flow through the ICA and VA was consistently ∼20-25% lower in KE (Fig. 4c-d, resp.). The KE-induced decrease in cerebral blood flow resulted from a drop in blood velocity and a trend towards lower arterial diameters (Table S1). This KE-induced drop in cerebral blood flow caused total cerebral oxygen delivery to be consistently lower in KE *vs.* CON (Fig. 4e). Similar to the cerebral oxygen status, skeletal muscle oxygenation index also increased from 15min to 4h in KE, while remaining stable in CON (Fig 4f).

**Figure 4.**
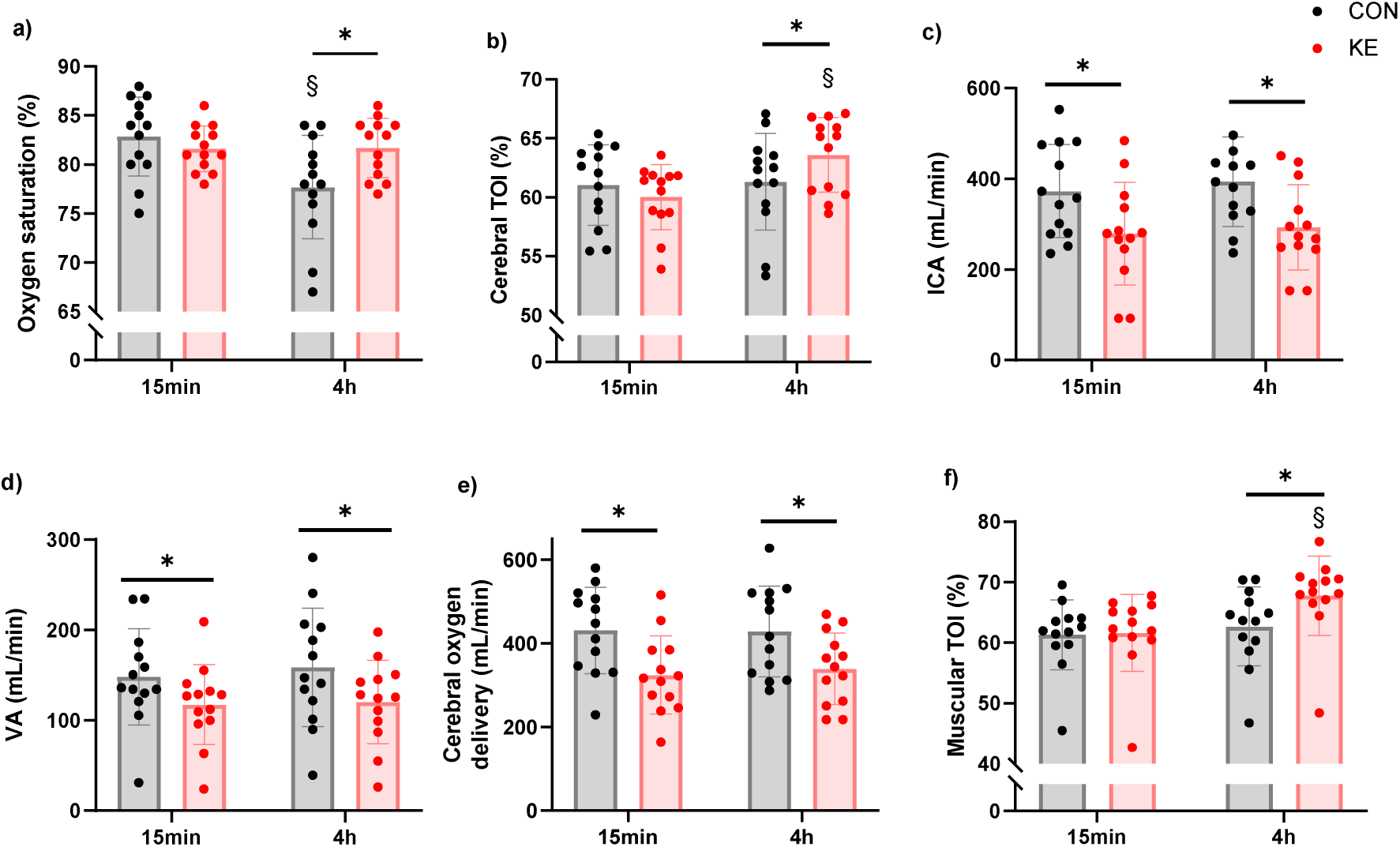
Mean (bar plots) ± SD (whiskers), as well as individual values for (a) arterial oxygen saturation, (b) cerebral (prefrontal cortex) tissue oxygenation index (TOI), (c) blood flow in the internal carotid artery (ICA), (d) blood flow in the vertebral artery (VA), and (e) cerebral oxygen delivery via the ICA and VA, and (f) muscular TOI. Measurements were performed after 15min and 4h of a 29h hypoxic protocol involving intermittent ketone ester (KE, red) or placebo (CON, black) ingestion. Data are shown for n = 13. No data were included for the participant that developed severe AMS after 1h in CON. *, p < 0.05 between KE and CON; §, p < 0.05 *vs.* 15min for indicated condition.

### KE shifted the autonomic nervous system towards increased sympathetic activity

KE consistently increased resting heart rate (Fig. 5a), while reducing the percentage of adjacent NN intervals that differ by more than 50 ms (pNN50, Fig. 5b), the root mean square of successive differences (RMSSD, Fig. 5c), as well as power of the high-frequency band (HF, Fig. 5d) which are all indicative of a shift towards increased sympathetic activity. This shift occurred without alterations in blood pressure (Table S2).

**Figure 5.**
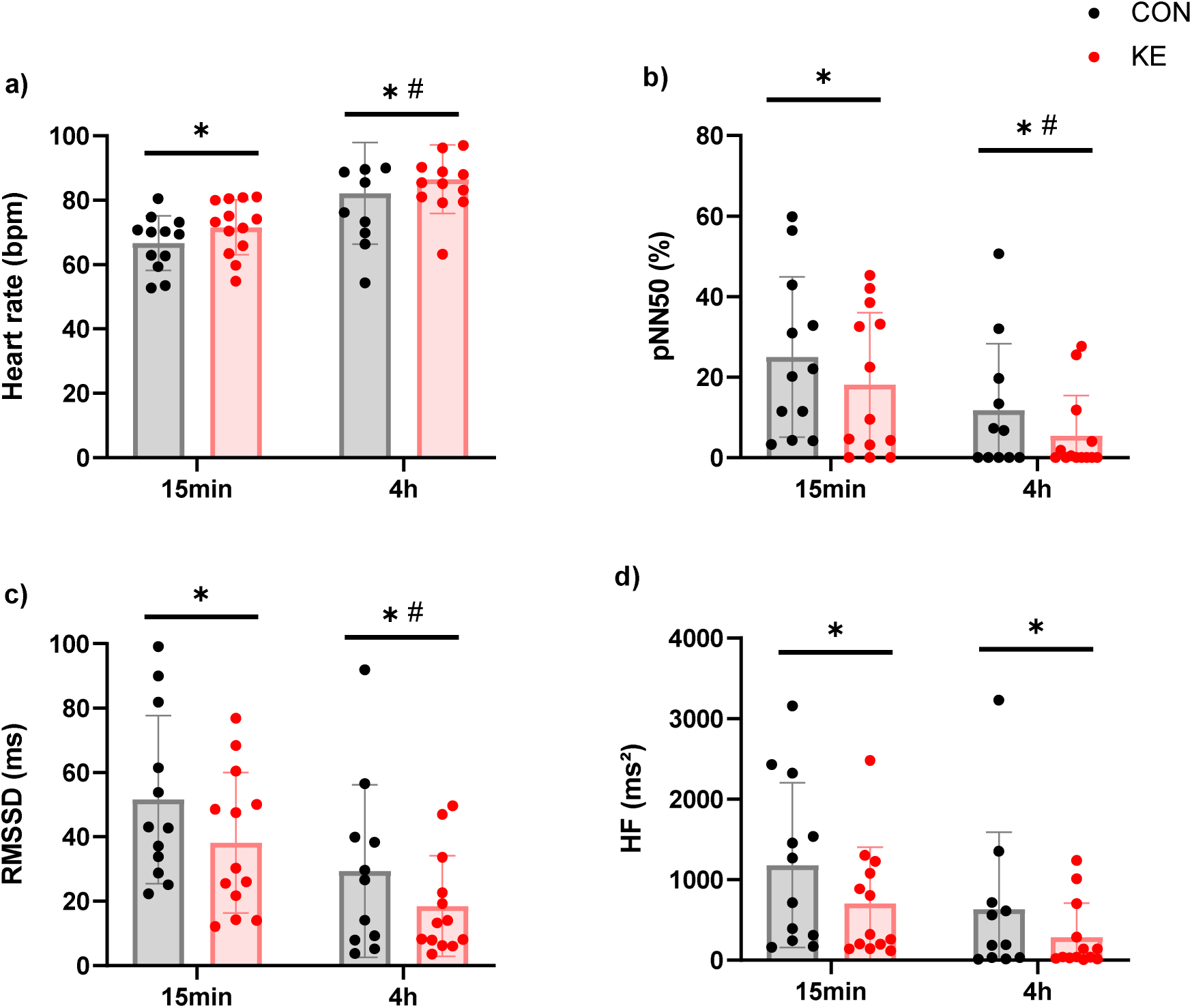
Mean (bar plots) ± SD (whiskers), as well as individual values for (a) heart rate, (b) the percentage of adjacent NN intervals that differ by more than 50 ms (pNN50), (c) the root mean square of successive differences (RMSSD), and (d) the power of the high-frequency band (HF). Measurements were performed after 15min and 4h of a 29h hypoxic protocol involving intermittent ketone ester (KE, red) or placebo (CON, black) ingestion. Data are shown for n = 13. No data were included for the participant that developed severe AMS after 1h in CON. *, p < 0.05 between KE and CON; #, p < 0.05 *vs.* 15min for both conditions.

### KE increased the hypoxic ventilatory response and altered acid-base balance

The improved hypoxemia may not only result from a reduction in tissue oxygen demand [20], but can also result from an increased ventilatory response [16]. In this perspective, KE consistently elevated ventilation (V̇_E_), as well as increased oxygen uptake rate (V̇O_2_) after 15min in hypoxia (Table 2 and Table S3). Yet between 15min and 4h, V̇O_2_ increased in CON but not in KE, resulting in similar values at 4h. Acid-base balance and capillary blood gasses are shown in Table 3. Compared to CON, KE consistently decreased blood pH, [HCO_3_^-^] and pCO_2_, while slightly increasing p50. Conversely, blood pO_2_ decreased to a similar extent in both conditions after 1.5h in hypoxia but was higher in KE *vs.* CON after 3h.

**Table 2.** Effect of ketone ester (KE) vs placebo (CON) ingestion on respiratory gas exchange parameters.

|  | CON | KE |
| --- | --- | --- |
| **$\dot{V}_E$ (L.min<sup>-1</sup>)** |  |  |
| 15min | 11.9 ± 1.6 | 12.9 ± 1.6 * |
| 4h | 13.4 ± 1.8 # | 14.7 ± 1.2 *# |
| **$\dot{V}O_2$ (L.min<sup>-1</sup>)** |  |  |
| 15min | 0.34 ± 0.04 | 0.37 ± 0.06 * |
| 4h | 0.36 ± 0.06 § | 0.35 ± 0.04 |
| **$\dot{V}CO_2$ (L.min<sup>-1</sup>)** |  |  |
| 15min | 0.35 ± 0.04 | 0.37 ± 0.05 |
| 4h | 0.31 ± 0.05 # | 0.29 ± 0.03 # |

Respiration was evaluated after 15min and 4h of a 29h hypoxic protocol in participants receiving either ketone ester (KE) or placebo (CON) drinks. $\dot{V}_E$ , ventilation; $\dot{V}O_2$ , oxygen consumption; $\dot{V}CO_2$ , carbon dioxide production. Data are means ± SD (n = 13). No data were included for the participant that developed severe AMS after 1h in CON. \*, p < 0.05 vs. CON; #, p < 0.05 vs. 15min in both conditions; §, p < 0.05 vs. 15min for indicated condition.

**Table 3.** Effect of ketone ester (KE) vs placebo (CON) ingestion on blood gasses and acid-base balance.

|  | CON | KE |
| --- | --- | --- |
| **pH** |  |  |
| Baseline | 7.414 ± 0.019 | 7.421 ± 0.012 |
| 1.5h | 7.432 ± 0.017 § | 7.395 ± 0.022 *§ |
| 3h | 7.450 ± 0.018 § | 7.392 ± 0.025 *§ |
| **[HCO<sub>3</sub><sup>-</sup>] (mM)** |  |  |
| Baseline | 25.7 ± 1.1 | 26.0 ± 0.8 |
| 1.5h | 25.8 ± 0.9 | 22.7 ± 1.0 *§ |
| 3h | 25.3 ± 1.1 | 21.0 ± 1.5 *§ |
| **pCO<sub>2</sub> (mmHg)** |  |  |
| Baseline | 41.3 ± 1.8 | 40.7 ± 1.3 |
| 1.5h | 40.0 ± 2.7 | 37.6 ± 1.9 *§ |
| 3h | 36.3 ± 1.8 # | 33.2 ± 1.8 *# |
| **p50 (mmHg)** |  |  |
| Baseline | 25.13 ± 1.57 | 24.74 ± 1.30 |
| 1.5h | 25.31 ± 1.00 | 26.32 ± 0.97 *§ |
| 3h | 24.68 ± 0.99 | 26.34 ± 0.99 *§ |
| **pO<sub>2</sub> (mmHg)** |  |  |
| Baseline | 84.3 ± 8.3 | 84.6 ± 6.5 |
| 1.5h | 42.0 ± 2.7 # | 43.9 ± 3.0 # |
| 3h | 40.9 ± 3.0 # | 46.3 ± 3.2 *# |

Blood gasses and acid-base balance were evaluated in capillary blood samples. Samples were obtained before hypoxic entry (baseline), and after 1.5 and 3h of a 29h hypoxic protocol involving intermittent ketone ester (KE) or placebo (CON) ingestion. Data are means ± SD (n = 13). No data were included for the participant that developed severe AMS after 1h in CON. \*, p < 0.05 vs. CON; #, p < 0.05 vs. baseline in both conditions; §, p < 0.05 vs. baseline for indicated condition.

## DISCUSSION

The present work sought to investigate whether increasing blood KB can improve hypoxic tolerance in humans and thereby alleviate the development/severity of AMS. For this purpose, participants underwent a 29h protocol twice at a simulated altitude of 4,000-4,500m involving intermittent exercise bouts, while receiving either KE or CON. Our results indicate that KE increased tolerated duration of simulated high-altitude exposure and reduced AMS symptoms. This was accompanied by an attenuation of the progressive decrement in arterial oxygen saturation during hypoxic exposure, and an increase in both cerebral and muscular oxygenation status. These observations were associated with a KE-induced increase in both ventilation and pO_2_, together with a reduction in cerebral oxygen delivery suggesting that KE likely reduced cerebral oxygen consumption. Furthermore, KE consistently reduced cerebral blood flow and shifted the autonomic nervous system towards increased sympathetic activity, which may both underly the observed attenuation of AMS.

A high degree of inter-individual variability in AMS susceptibility is well established. In this regard, the normobaric hypoxic protocol elicited AMS in 64% (9 out of 14) of the individuals in the CON condition, resulting in an average tolerated duration of ∼15h. This approximates the expected incidence at terrestrial altitude given earlier studies reporting an incidence rate of 53% in mountaineers at 4,243m [3] and 4,559m altitude [21]. Interestingly, KE ingestion completely negated the development of AMS in 2 individuals, as well as increased hypoxic tolerance by 32% on average and by 99% in AMS-developing participants.

The primary trigger for AMS is a hypoxia-induced drop in arterial oxygen saturation. This is clearly exemplified by the nearly immediate recovery from AMS upon administration of oxygen or hyperbaria [22], and the direct relationship between the extent of hypoxemia and AMS incidence and severity [6]. In our study, SpO_2_ values dropped to ∼82% after 15min of hypoxic exposure in both conditions, which is in perfect agreement with the expected SpO_2_ at the given altitude [23]. By the 4h timepoint, SpO_2_ values had dropped by an additional 5% in CON, which most likely reflects an inability to cope with the hypoxic stress [9]. Remarkably, this final drop in oxygen saturation was fully negated by KE supplementation.

This KE-induced attenuation of oxygen desaturation is in line with the results from an earlier study by our research group [16]. In this study, KE negated the drop in oxygen saturation during the final ∼30min of a 3h submaximal cycling bout wherein simulated altitude gradually increased from 1,000m to 3,000m. Interestingly, both in our earlier work as well as in the current study, SpO_2_ values were increased by KE relative to CON only after a few hours of hypoxic/ketone exposure in combination with exercise (*e.g.,* ∼2.5-4h), but not during the earlier timepoints (*e.g.,* ∼15min-2h). The extent of altitude/hypoxemia was more pronounced after 15min in the current study than after 2h in the earlier study. This suggests that the ability of KE to raise blood oxygen saturation is independent of the extent of altitude/hypoxemia, but rather requires a given period of (hypoxic) time or exercise load. This time-dependent effect may for instance be linked to the levels of glucagon as earlier data showed that simultaneous administration of glucagon and ßHB, but not glucagon nor ßHB alone, doubled hypoxic survival time in mice [13]. In this regard, plasma glucagon levels increased by 25% after 5h in hypoxia. Another explanation for the delayed improvement in oxygenation with KE might be related to the previously demonstrated KB-associated facilitation of mitochondrial respiration, occurring only after prolonged KB exposure [24].

Despite a consistently lower cerebral oxygen delivery in KE, cerebral oxygenation was similar between both conditions at the 15min timepoint. This supposedly indicated that KE lowered oxygen demand or consumption by the brain, potentially indicating improved cerebral oxygen efficiency. However this did not coincide with changes in SpO_2_ or pO_2_, which were similar between KE and CON at 15min. This was probably explained by the consistent ketosis-induced hyperventilation which, in line with previous research [16, 25], concomitantly provoked a higher V̇O_2_ in KE compared to CON and thus fully compensated the lowered cerebral oxygen demand. The observed, delayed attenuation of arterial oxygen desaturation with KE can either result from increased arterial pO2 or a left-shift of the oxyhemoglobin dissociation curve. Our results indicate that the former mechanism was at play given that the KE-induced acidosis caused a right-shift of the curve (increased p50), while capillary pO2 values were higher in KE *vs.* CON after 3h. It should be highlighted that pO2 values were obtained using a capillary sampling method, which might underestimate actual arterial pO2. Nevertheless, these data confirm earlier observations showing an increase in arterial, capillary and hippocampal oxygenation [16, 26, 27] upon endogenous and exogenous ketosis. In turn, an increased pO2 can either result from a higher oxygen uptake through increased ventilation or from a decreased cellular oxygen consumption. After prolonged hypoxic exposure, a gradually augmented hypoxic ventilatory response increased V̇O_2_ in CON, which likely promoted the observed drop in oxygen saturation from 15min to 4h. Despite a slight additional increase in hyperventilation in KE, oxygen consumption did not further increase and SpO_2_ values remained stable. Interestingly, KE intake increased cerebral oxygenation after 4h, even in combination with the before mentioned lower cerebral oxygen delivery, suggesting a potential higher increase in cerebral oxygen efficiency after prolonged exposure. Such effect may have also been present in other tissues given the observed KE-induced increase in oxygenation status of the *m. vastus lateralis*, and supports earlier *ex-vivo* data indicating that KB can increase mechanical work to oxygen ratio [28].

Hypoxemia is considered to evoke AMS via multiple mechanisms, with recent evidence indicating that the specific pathophysiological mechanism depends on the time course by which AMS develops [9]. In this perspective, AMS development during the first 29h is mostly caused by an increase in (i) relative parasympathetic activity, and (ii) cerebral blood flow. Both parasympathetic contribution, as evidenced by resting heart rate, and the HRV indices pNN50, RMSSD, and HF power, as well as blood flow in both the ICA and VA, the main arteries for intracranial blood supply, were markedly reduced upon KE. This suggests that KE may have lowered AMS symptoms via both mechanisms. An increase in sympathetic drive has also been reported in some, but not all, earlier studies looking at the acute effect of ketosis on sympathetic activity in normoxia [29–31], and may a.o. result from a KE-induced suppression of atrial natriuretic peptide [31]. This suggests that the observed increase in sympathetic tone in our study likely directly results from ketosis as the KE-induced attenuation of hypoxemia would rather attenuate sympathetic stimulation. In contrast, cerebral blood flow has consistently been shown to improve during acute ketosis under normoxic conditions both in rats [32], and in healthy and obese adults [33, 34]. This suggests that the KE-induced reduction in cerebral blood flow observed in our study was not a primary effect of ketosis, but occurred as a secondary effect mediated by the increase in oxygen saturation upon KE.

Collectively, these data indicate that KE ingestion may evolve as a novel, non-pharmacological intervention to increase hypoxic tolerance and alleviate AMS. Although a gradual ascent is still the best strategy to prevent AMS [35], the carbonic anhydrase inhibitor acetazolamide (AZ) is currently considered as the preferred drug to reduce AMS [11]. Interestingly, the mechanism by which AZ increases hypoxic tolerance shows some analogy with the observed physiological effects of KE. Similar to AZ-induced inhibition of carbonic anhydrase (CA) [36], βHB has been shown to exert an inhibitory effect on human CA activity [37] and therefore might provoke a similar physiological response. Administration of AZ causes a bicarbonate diuresis thereby generating a metabolic acidosis which via an increased ventilatory drive results in better arterial oxygenation [10]. Nevertheless, there are also some profound differences given that AZ for instance induces diuresis at high dosages [38], while KE is rather anti-diuretic [31]. It would be inappropriate to compare the effectiveness of AZ with KE given the limited sample size of our study. Nevertheless, the observed increase in arterial oxygen saturation upon KE (∼+4%) was at least similar as typically seen following AZ (∼+2 to 5%) at comparable altitudes [39, 40].

Despite providing novel insights, we would like to acknowledge a few limitations of the present study. In most cases, AMS manifests within the timeframe of the study (*e.g.,* within 29h following acute altitude exposure). But, in some participants symptom severity peaks at a slightly later timepoint, particularly triggered by subclinical pulmonary edema [9]. As such, it remains to be determined if KE can also prove beneficial for this AMS type. Second, it should be highlighted that our study was performed in normobaric hypoxia. Earlier research clearly showed that the response to normobaric *vs.* hypobaric hypoxia elicits comparable physiological effects [41, 42], yet also minor differences exist such as slightly higher AMS scores in hypobaric hypoxia [43]. Along a similar line, it remains to be identified if similar effects would also be observed in women, who have inconsistently been shown to have a potential higher incidence of AMS [2, 44]. AMS development is also inversely related to energy intake [45]. In this perspective, our placebo drink contained less calories compared to the KE drink. But it is unlikely that this may have provoked the observed effect given that all participants received a caloric surplus, and given that lower energy intake is rather a consequence than a cause of AMS [46].

In conclusion, our data are the first to indicate that exogenous ketosis improves hypoxic tolerance and attenuates the development of AMS. This is mediated by an improved arterial oxygenation in combination with increased cerebral and muscular oxygenation, lower cerebral blood flow, and increased sympathetic dominance. The increase in cerebral oxygenation occurred along with a reduction in cerebral oxygen supply, thereby providing the first *in-vivo* human data to indicate that ketosis can lower cerebral oxygen demand at high altitude.

## Data Availability

All data produced in the present study are available upon reasonable request to the authors

## ADDITIONAL INFORMATION

### Author contributions

All experiments were performed within the Exercise Physiology Research Group and the Bakala Academy-Athletic Performance Center at the KU Leuven, Belgium. Conception and design of the study: MS, TD and CP. Data collection and/or data analyses: MS, DT, WL, RR, TD and CP. Interpretation of the data and manuscript drafting: MS and CP. All authors critically revised the manuscript and approved the final version of the manuscript.

## Acknowledgements

The authors wish to thank all participants for their dedicated cooperation in this study. We also thank Ms. Monique Ramaekers for skillful assistance during the experimental trials.

