## Supplemental tables and figures for "Exogenous ketosis attenuates acute mountain sickness and mitigates high-altitude hypoxemia"

**Summary:** Ketone ester intake attenuated the development of acute mountain sickness at a simulated altitude of 4,000-4,500m. This likely resulted from a mitigation of arterial and cerebral hypoxemia, reduced cerebral blood flow and increased sympathetic drive.

**TABLES**

**Table S1.** Effect of ketone ester (KE) vs placebo (CON) ingestion on blood flow velocity and vessel diameter.

| | **CON** | **KE** |
|---|---|---|
| **$v_{ICA}$ (cm.s$^{-1}$)** | | |
| 15min | 23.37 ± 3.94 | 21.08 ± 3.29 * |
| 4h | 23.12 ± 2.05 | 20.23 ± 2.56 * |
| **$v_{VA}$ (cm.s$^{-1}$)** | | |
| 15min | 18.74 ± 3.68 | 15.51 ± 4.60 * |
| 4h | 18.58 ± 3.62 | 15.24 ± 2.89 * |
| **$d_{ICA}$ (cm)** | | |
| 15min | 0.57 ± 0.07 | 0.52 ± 0.10 |
| 4h | 0.60 ± 0.08 | 0.55 ± 0.10 |
| **$d_{VA}$ (cm)** | | |
| 15min | 0.39 ± 0.05 | 0.38 ± 0.05 |
| 4h | 0.40 ± 0.06 | 0.39 ± 0.06 |

Blood velocity (v) and diameter (d) of the internal carotid artery (ICA) and vertebral artery (VA) used to calculate cerebral blood flow. Measurements were performed after 15min and 4h of a 29h hypoxic protocol involving intermittent ketone ester (KE) or placebo (CON) ingestion. Data are means ± SD (n = 13). No data were included for the participant that developed severe AMS after 1h in CON. *, $p < 0.05$ *vs.* CON. Main effect of group: $p = 0.0908$ and $p = 0.0637$ for $d_{ICA}$ and dva, respectively.

**Table S2.** Effect of ketone ester (KE) vs placebo (CON) ingestion on blood pressure.

| | CON | KE |
|---|---|---|
| **SBP (mmHg)** | | |
| Baseline | 123 ± 9 | 123 ± 7 |
| 15min | 127 ± 9 # | 128 ± 6 # |
| 4h | 117 ± 7 # | 118 ± 10 # |
| **DBP (mmHg)** | | |
| Baseline | 71 ± 10 | 72 ± 9 |
| 15min | 71 ± 9 | 72 ± 7 |
| 4h | 70 ± 8 | 71 ± 6 |
| **MAP (mmHg)** | | |
| Baseline | 88 ± 9 | 89 ± 7 |
| 15min | 89 ± 8 | 90 ± 7 |
| 4h | 86 ± 7 | 86 ± 7 |

Mean arterial blood pressure (MAP) was evaluated before hypoxic entry (baseline), as well after 15min and 4h in hypoxia. MAP was calculated from systolic blood pressure (SBP) and diastolic blood pressure (DBP). Data are means ± SD (n = 13). No data were included for the participant that developed severe AMS after 1h in CON. #, $p < 0.05$ *vs.* baseline for both conditions.

**Table S3.** Effect of ketone ester (KE) vs placebo (CON) ingestion on respiratory gas exchange parameters.

| | CON | KE |
|---|---|---|
| **BF ($min^{-1}$)** | | |
| 15min | 14 ± 4 | 14 ± 4 |
| 4h | 17 ± 3 § | 18 ± 3 § |
| **TV (L)** | | |
| 15min | 0.87 ± 0.23 | 0.97 ± 0.40 |
| 4h | 0.80 ± 0.15 | 0.82 ± 0.12 |
| **$F_I$ (%)** | | |
| 15min | 44 ± 2 | 44 ± 3 * |
| 4h | 44 ± 3 | 47 ± 2 *§ |

Respiration was evaluated after 15min and 4h of a 29h hypoxic protocol in participants receiving either ketone ester (KE) or placebo (CON) drinks. BF, breathing frequency; TV, tidal volume; $F_I$, fractional inspiration time. Data are means ± SD (n = 13). No data were included for the participant that developed severe AMS after 1h in CON. *, $p < 0.05$ *vs.* CON; §, $p < 0.05$ *vs.* 15min for indicated condition.

**FIGURES**

**Figure S1.** Mean (bar plots) ± SD (whiskers) as well as individual values for plasma glucagon concentrations. Measurements were performed before breakfast, still in normoxia (baseline) and after 5h of a 29h hypoxic protocol involving intermittent ketone ester (KE, red) or placebo (CON, black) ingestion. Data are shown for n = 13. No data were included for the participant that developed severe AMS after 1h in CON. #, $p < 0.05$ *vs.* baseline for both conditions.

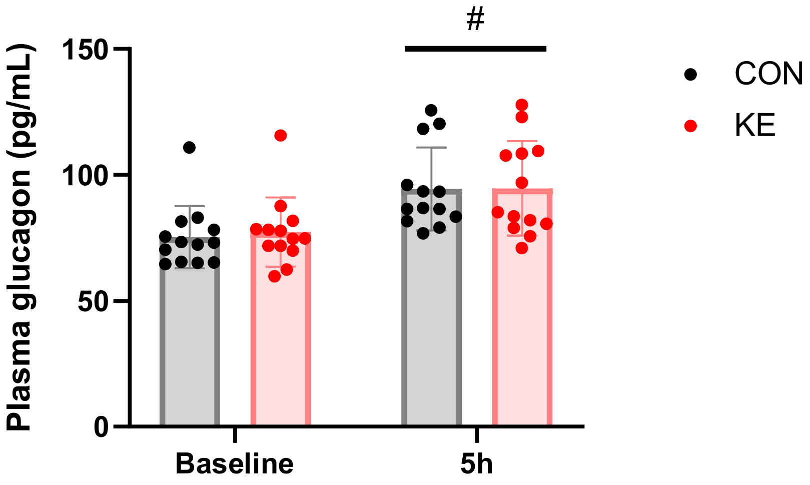
