## Supplemental information for "Exogenous ketosis attenuates acute mountain sickness and mitigates high-altitude hypoxemia"

**Summary:** Ketone ester intake attenuated the development of acute mountain sickness at a simulated altitude of 4,000-4,500m. This likely resulted from a mitigation of arterial and cerebral hypoxemia, reduced cerebral blood flow and increased sympathetic drive.

**SUPPLEMENTARY INFORMATION**

*Exercise bouts.* During the familiarization session, participants performed an incremental test to determine their maximal oxygen uptake rate ($\dot{V}O_2max$) on a cycling ergometer (Avantronic Cyclus II, Leipzig, Germany). After 15min of warming up at 70W, resistance was set at 100 W and increased by 25 W/30sec until voluntary exhaustion. The protocol included four 30min submaximal exercise bouts (30min at 1.5 $W.kg^{-1}$; performed after 3, 6, 7 and 8h at altitude), 2 maximal exercise bouts (10min at 1.5 $W.kg^{-1}$ followed by 100 W + 25 W/30sec until voluntary exhaustion; performed after 1.5 and 25h at altitude), and one combined exercise bout (30min at 1.5 $W.kg^{-1}$ followed by 100 W + 25 W/30sec until voluntary exhaustion; performed after 26.5h at altitude).

*Blood and tissue (prefrontal cortex and skeletal muscle) oxygenation status.* Blood oxygen saturation ($SpO_2$) was measured after 15min and 4h in hypoxia, at 2 Hz using a pulse oximeter (Nellcor PM10N, Medtronic, Minneapolis, USA) with an infrared sensor placed ~2 cm above the left eyebrow. Values were recorded after 10min of bedrest in supine position and data are presented as the average value of the last 30sec. Cerebral and skeletal muscle oxygenation status were assessed by analysis of tissue oxygenation index (TOI) using near infrared spectroscopy (NIRS), also after 15min and 4h in hypoxia. The probes of a NIRO-200 spectrometer (Hamamatsu, Japan) were attached ~2 cm above the right eyebrow for cerebral oxygenation and centrally on the belly of the right *m. vastus lateralis* for skeletal muscle oxygenation. In order to maintain a fixed interoptode distance of 4 cm, crucial to ensure a constant penetration depth of ~2 cm into the muscle/brain tissue, the emitter and detector probes were inserted in a dark-colored rubber spacer. These spacers were attached to the participants using an elastic non-transparent bandage and double-sided adhesive tape to prevent displacement or interference from external light. Before each experimental session, the involved skin was shaved and cleaned to exclude any signal disturbance by hair or impurities. Moreover, the contour lines of the rubber spacer were marked on the skin with a dermatological pen during preparation for the first session, to ensure identical positioning for every measurement. Participants were asked to preserve and refresh these marks during the washout period in order to maintain this position during the second session. After initial data collection, NIRS data were preprocessed (Matlab R2023a, The Mathworks, Natick, MA) using a fourth-order Butterworth filter with a cut-off frequency of 0.05 Hz [16]. Data were analyzed over 1-min long time chunks.

*Cerebral blood flow.* Vessel diameter (d) and blood velocity (v) of the internal carotid artery (ICA) and the vertebral artery (VA) were assessed by a trained and experienced ultrasonographer after 15min and 4h using duplex ultrasonography (Vivid E9, EG Healthcare, New York, USA, with a 9L linear transducer of 2.4 - 10.0 MHz). Measurements were performed in agreement with the technical recommendations as described by Thomas et al. [47]. Artery diameters were measured in the sagittal axes using B-mode imaging, while blood velocity was assessed using pulse-wave mode for later offline analysis. Vessel location and sample volume were determined on an individual basis during the familiarization session, with careful consideration of the diagnostic details of the ICA, and replicated for every measurement. Moreover, the Doppler approach angle was set at 60°, yet angle corrections were applied for tortuous or oblique-angled vessels. Gain and dynamic range were fixed during the familiarization session and unaltered throughout the entire study period. The ICA was assessed at a distance of at least 1.5 cm from the carotid bifurcation in order to guarantee reproducibility and to avoid turbulence. The VA was analyzed between C4 and C5, or alternatively C5 and C6, however the exact location was replicated for each measurement within each participant. Vessel diameters of the ICA and VA ($d_{ICA}$ and $d_{VA}$, resp.) were measured over 30 sec and 5 consecutive cardiac cycles were included for analysis. Blood velocity ($v_{ICA}$ and $v_{VA}$, resp.) recordings were collected over 60 sec and analyzed for beat-to-beat TAMEAN using the available ultrasound review software EchoPAC (GE Healthcare, New York, USA). Blood flow ($Q_{ICA}$ and $Q_{VA}$, resp.) was calculated as follows: $Q_i = v_i * \left(\frac{d_i}{2}\right)^2 * \pi * 60$ with i being either ICA or VA.

*Respiratory gas exchange measurements.* Indirect calorimetry (Cortex Metalyzer IIIb, Leipzig, Germany) was used during the resting measurement bouts after 15min and 4h in hypoxia to measure breath-by-breath gas exchange data [*i.e.,* minute ventilation ($\dot{V}E$), oxygen uptake ($\dot{V}O_2$) and carbon dioxide production ($\dot{V}CO_2$)], as well as breathing patterns [*i.e.*, breathing frequency (BF) and tidal volume (TV)]. Data collection started after 10min of bedrest to ensure physiological homeostasis, and data are presented as the average values of the last 5 min.

*Blood pressure and heart rate.* Resting heart rate and RR intervals (Polar H10, Polar, Kempele, Finland) were measured after 15min and 4h in hypoxia. Data were collected after 10min of bedrest, and the data are presented as the average values of the last minute. Heart rate variability (HRV) parameters were assessed using Kubios (Kubios HRV Standard 3.5.0, Kubios Oy, Kuopio, Finland). Blood pressure (Omron M6, Omron healthcare, Kyoto, Japan) was measured before hypoxic entry, as well as after 15min and 4h in hypoxia, on the left arm in sitting position. Systolic (SBP) and diastolic (DBP) blood pressure were used to calculate the mean arterial pressure [MAP = $DBP + \frac{1}{3}(SBP - DBP)$].

*Venous blood samples.* Before hypoxic entry (baseline) and after 5h in the hypoxic protocol, venous blood samples were obtained from an antecubital vein (Venoject, Terumo, Tokyo, Japan) and collected into vacuum tubes containing EDTA (Becton Dickinson (BD) Vacutainer, Eysins, Switzerland). Tubes were centrifuged (1500 rpm for 10 min at 4°C) and the supernatant was stored at -80°C until later analysis. A commercially available enzyme-linked immunosorbent assay (ELISA) was performed to determine plasma glucagon levels (DGCG0, R&D Systems, Minneapolis, MN, USA).

*Capillary blood samples and analyses.* Immediately before and 30min after each supplement, and upon waking up, capillary blood samples were obtained from a hyperaemic earlobe for immediate determination of D-ß-hydroxybutyrate (GlucoMen Areo 2K-meter with ß-ketone sensor strips, A. Menarini Diagnostics, Firenze, Italy ). In addition, 70 µl capillary blood was collected from a hyperemic earlobe into a capillary tube (safeCLINITUBE, Radiometer Medical ApS, Copenhagen, Denmark) (i) before the first supplement, in normoxia, as well as after (ii) 1.5 and (iii) 3h in hypoxia, in resting conditions. After immediate mixing for 10sec, samples were analyzed for acid-base balance, $pO_2$, and $pCO_2$ (ABL90 FLEX analyzer, Radiometer Medical ApS, Copenhagen, Denmark).

*Dietary control.* Participants were provided, in sequential order (i) a carbohydrate-rich dinner (~5,600 kJ; 69% carbohydrate, 16% fat, 15% protein) the evening before the experimental session, (ii) a breakfast (~4,100 kJ; 64% carbohydrate, 26% fat, and 10% protein) 30min before entering the hypoxic room, (iii) a lunch after 3.5h in hypoxia (~3,950 kJ; 53% carbohydrate, 26% fat, and 18% protein), (iv) a light evening meal after 10.5h in hypoxia (~2,330 kJ; 67% carbohydrate, 5% fat, and 26% protein), and (v) an identical breakfast on the same time of the day as on day 1. In addition, a carbohydrate-rich snack (~660 kJ; 92% carbohydrate, 2% fat, and 4% protein) was provided after 2.5, 6.5, 9, 12 and 26.5 h in hypoxia. Participants were allowed to ask for more food during the first session if they felt hungry, and additional caloric intake was then replicated during the second session.

*Hypoxic facility.* The fully automated normobaric hypoxic system (b-cat, No. 3113205, Van Amerongen CA Technology, Tiel, The Netherlands) in Bakala Academy (Leuven, Belgium) was programmed to achieve the preset $F_IO_2$ levels while maintaining low $F_iCO_2$ (<0.05%) levels during the full duration of the study. The electrochemical $O_2$ and $CO_2$ sensors of the system were calibrated on a daily basis with a 2.5% $CO_2$-97.5% $N_2$ gas mixture (Air Liquide, stad en land) and fresh, outside air. Moreover, before every hypoxic exercise or resting measurement, $F_IO_2$ was verified using a portable oxygen sensor (G0X 100T, Greisinger, Regenstauf, Germany). Ambient air temperature (19 °C), relative humidity (60%) and air ventilation were also standardized.

*Randomization.* Allocation of the subjects to the experimental conditions was performed *at random*, yet stratified based on oxygen saturation after 30min at a simulated altitude of 4,000m. Recruitment was performed in October 2022, and experimental sessions in November-December 2022. Subjects and researchers were blinded to the experimental conditions by using a placebo for KE similar in taste and appearance. At the level of the investigators, blinding was ensured by having the randomization done by a person who was otherwise not involved in the experimental testing. Moreover, the researchers performing the measurements were unaware of the randomization method (e.g. stratified randomization), and blood ketone concentrations were assessed by an investigator who was neither involved in the randomization procedure nor in any of the other measurements. To maintain the blinding, nowhere in the study the test products and corresponding placebos were revealed and identity concealment codes were used in the randomization list.
